# A new rescue assay for genetic diagnosis of oculocutaneous albinism using MNT1 knock-out cells

**DOI:** 10.1101/2025.11.26.25341047

**Authors:** Elina Mercier, Vincent Michaud, Angèle Sequeira, Benoit Arveiler, Sophie Javerzat

## Abstract

The molecular diagnosis of albinism is hampered by a significant number of genetic variants of unknown significance (VUS) including a majority of missense and in-frame insertion deletion variants. This contributes to the high rate of unresolved genetic diagnosis for this disease. We designed a straightforward test of missense VUS in albinism genes based on functional rescue. As a proof of concept, the assay was set up for testing variants in the *TYR* gene associated with oculocutaneous albinism type 1. The *TYR* gene was knocked-out in the human melanogenic MNT1 cell line and the resulting unpigmented clones used as host cells for rescue experiments. Selected VUS and control sequences were run through the assay. Expression of tyrosinase was quantified by Western-blot, melanin synthesis was evaluated by direct observation as well as absorbance monitoring. One VUS, p.Ser270Phe (S270F) can be classified as pathogenic as it fails to restore pigmentation, whereas rescue was achieved with D305E and A391T. The two most frequent missense VUS of *TYR*, S192Y and R402Q, were also tested independently or in combination confirming the pathogenic effect of their association in *cis*. All in all, this new assay is straightforward enough to be transposed in diagnosis laboratories and can be considered for testing variants in other albinism genes such as *TYRP1* and *SLC45A2*.

## 1. Introduction

Albinism is a rare heterogeneous condition that is characterized by hypopigmentation of the skin, hair and/or eye and systematic vision alterations (1). Twenty-two genetic subtypes have been identified so far, with the most frequent form OCA1 (oculocutaneous type 1) accounting for over 40% of cases (2). OCA1 is associated with genetic variants in *TYR*. The *TYR* gene encodes the enzyme tyrosinase that triggers the initial steps in the melanogenic cascade (3). Patients with biallelic loss-of-function variants in *TYR* present with white skin and hair and severe visual impairment. By contrast, individuals with hypomorphic variants fall somewhere along a phenotypic spectrum depending on their *TYR* diplotype and the associated level of residual tyrosinase activity. We previously identified haplotypic combinations involving two frequent missense variants of *TYR* (c.575C>A/p.S192Y and c.1205G>A/p.R402Q) that are associated with clinically diagnosed albinism depending on a single nucleotide variant (SNV), c.-301C>T [rs4547091] located in the promoter (4). Although this statistical study was unable to test the controversial additive role of S192Y on the haplotype [c.-301C>T; c.1205G>A/p.R402Q], it challenged the classical paradigm which attributes causality to rare completely penetrant biallelic variants. It now serves as a guide for accurate genetic testing taking into account the quantitative effect of variants of unknown significance (VUS) according to the individual diplotype including [c-301C; c.1205G>A/p.R402Q] (5). Despite the recent development of artificial intelligence-based *in silico* prediction tools (6), 193 missense variants *TYR* are still reported as VUS in the ClinVar database (https://www.ncbi.nlm.nih.gov/clinvar). These variants, which are sometimes the only ones found in patients awaiting genetic diagnosis, justify the implementation of rapid and effective functional tests.

The activity of recombinant tyrosinase carrying amino acid substitutions corresponding to identified missense variants can be tested *in vitro* (7,8). One disadvantage of this strategy is that it relies on the use of high-level overexpression systems such as insect larvae which are not routinely available in molecular diagnostic laboratories. Furthermore, the assay does not assess the mutant protein’s ability to traffic across the endoplasmic reticulum, reach the melanosome and interact with the other enzymes TYRP1 and DCT to produce final melanin.

Here, we knocked-out the *TYR* gene in the human melanocytic MNT1 cell line and used the resulting clones to design a cell-culture based rescue assay. The MNT1 line was chosen because it harbors a functional melanogenic machinery unlike most melanoma derived cell lines. MNT1 cells are widely used for the study of pigmentation pathologies (9). Their morphology, transcriptome and melanosome content are similar to those of primary melanocytes (10).

Eight variants including three VUS as well as the two frequent variants S192Y and R402Q alone or in combination were tested for their ability to restore melanin production qualitatively and quantitatively. The assay is straightforward, quick and inexpensive to set-up and can be extrapolated to other albinism gene variants such as *TYRP1* (OCA3) and *SLC45A2* (OCA4).

## 2. Results

### 2.1 Production and characterization of *TYR-*KO clones derived from melanogenic MNT1 cells

As detailed in the Materials and Methods section, exon 2 of the *TYR* gene was targeted by CRISPR-Cas9 engineering in the human melanogenic MNT1 cell line. *TYR*-KO clones were positively screened according to their hypopigmented phenotype. Three independent clones, homozygotes for a frameshift mutation were selected and further characterized (Fig. 1). RT-qPCR dosage (Fig. 1a) indicated a near total loss of *TYR* mRNA confirming nonsense-mediated mRNA decay (NMD) due to the introduction of a premature stop codon (11). Western-blot experiments confirmed the absence of tyrosinase protein in each *TYR*-KO clone (Fig. 1b). Complete loss of tyrosinase abrogated melanin production as illustrated by direct observation of the cultured cells and cell pellets (Fig. 1c). Conventional dosage based on pigment solubilization followed by spectrometric measurement of absorbance at 490 nm can serve as a quick method to assess melanin production (Fig. 1d) as previously reviewed (12).

**Fig. 1:**
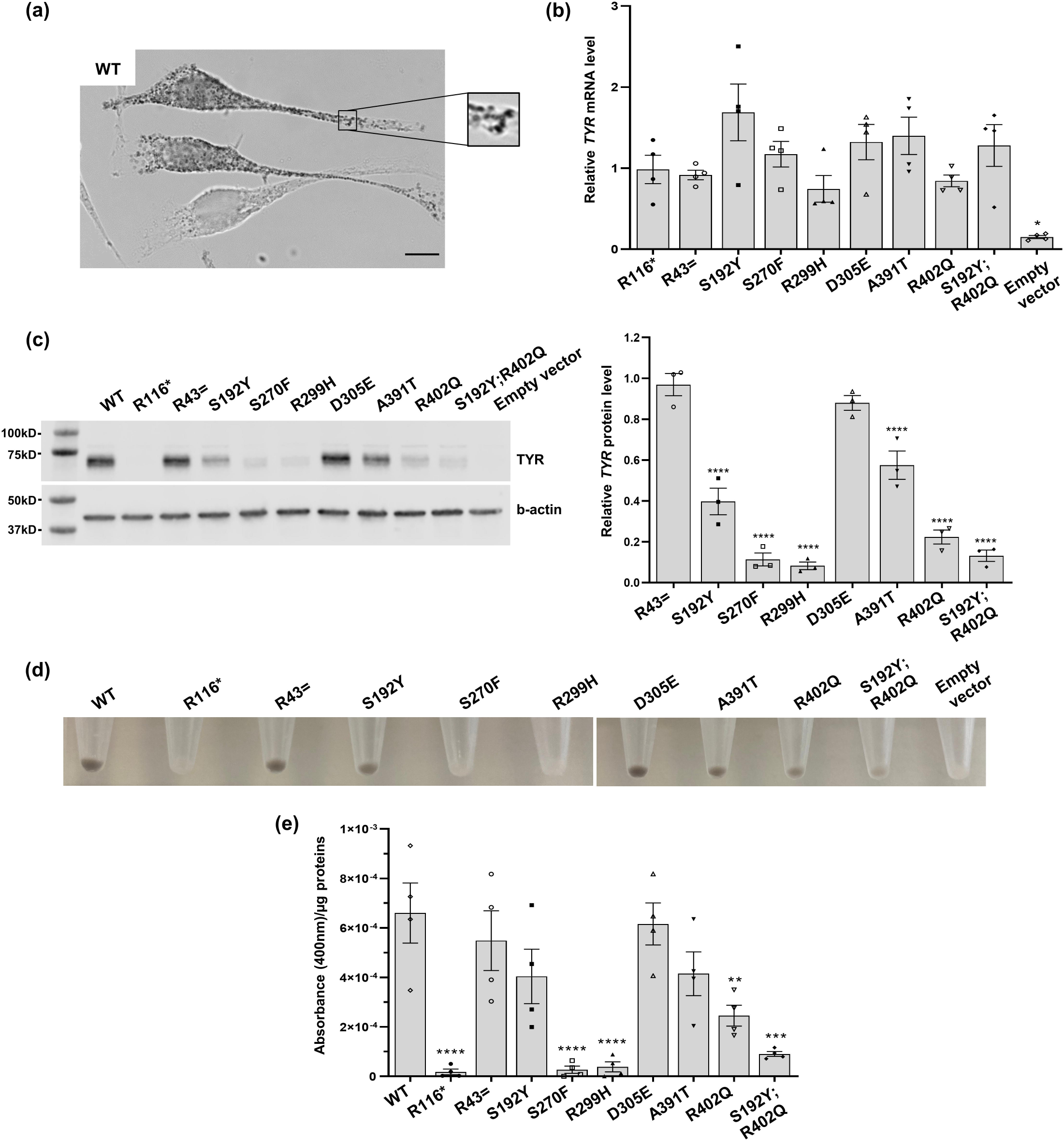
Characterization of selected *TYR*-KO MNT1 clones. Loss of function of *TYR* was confirmed with *TYR* expression monitoring, at the mRNA level by RT-qPCR (a) and at protein level by Western-blot (b). (a) *TYR* relative expression normalized to *B2M* reference gene expression in the three selected *TYR-*KO clones compared to wild-type MNT1 cells using the 2^−ΔΔCt^ method. Data are presented as mean ± SEM (n=3). Means are compared using a t-test, ** indicates p<0.01. (b) Representative image of Western-blot detection of tyrosinase confirming the absence of protein in *TYR*-KO clones. The loss of function of the *TYR* gene in MNT1 cells results in impairment of the cells’ ability to produce pigment, which can be directly observed from cultured cells (c) or evaluated with a melanin dosage (d). (c) Cell pigmentation can be assessed using contrast or bright field microscopy (upper and middle panel) or pictures of cell pellets (low panel). Microscope magnification x40. (d) Normalized melanin dosage was performed from samples corresponding to the three *TYR-*KO clones compared to MNT1 cells. Data are shown as mean ± SEM (MNT1 n=6, *TYR-*KO n=8). Means are compared using a t-test, **** indicates p<0.0001.

### 2.2 Selection of missense variants for evaluating the rescue assay

A total of eight variants of *TYR* were selected to construct the recombinant expression vectors (Table 1). Three variants with already established significant prediction scores were included to serve as controls: the synonymous variant R43= is predicted likely benign. The truncating variant R116* and the missense variant R299H are both reported as pathogenic in ClinVar and have been identified in several unrelated patients of our cohort with a 2^nd^ pathogenic variant in *trans*. Variant R299H was first reported in 1992 (13). Three rare missense VUS (S270F; D305E and A391T) were selected. Variant S270F is absent from the ClinVar database. It has been identified only once in our cohort in a heterozygous patient with multiple carcinomas but no signs of albinism. Variant D305E is reported 8 times as a VUS in ClinVar. Three South-Asian individuals are identified as homozygotes for this variant in gnomAD. In the literature, D305E has been reported twice in patients with albinism but no identified 2^nd^ *TYR* variant in *trans* (14,15). In our local cohort it was found twice as heterozygous in patients clinically diagnosed with albinism. A391T is reported once in the Clinvar database as VUS. It was found once in a heterozygote patient with no signs of albinism in our cohort. The two frequent variants S192Y and R402Q were also selected to be tested on their own or in *cis* one to the other independently of the c.-301C>T promoter SNV (16). Of note, the haplotype S192Y/R402Q is more frequent in Europe (f=0.01) than in the worldwide population (f=0.003) as estimated by analyzing the LDlink database (17).

**Table 1:**
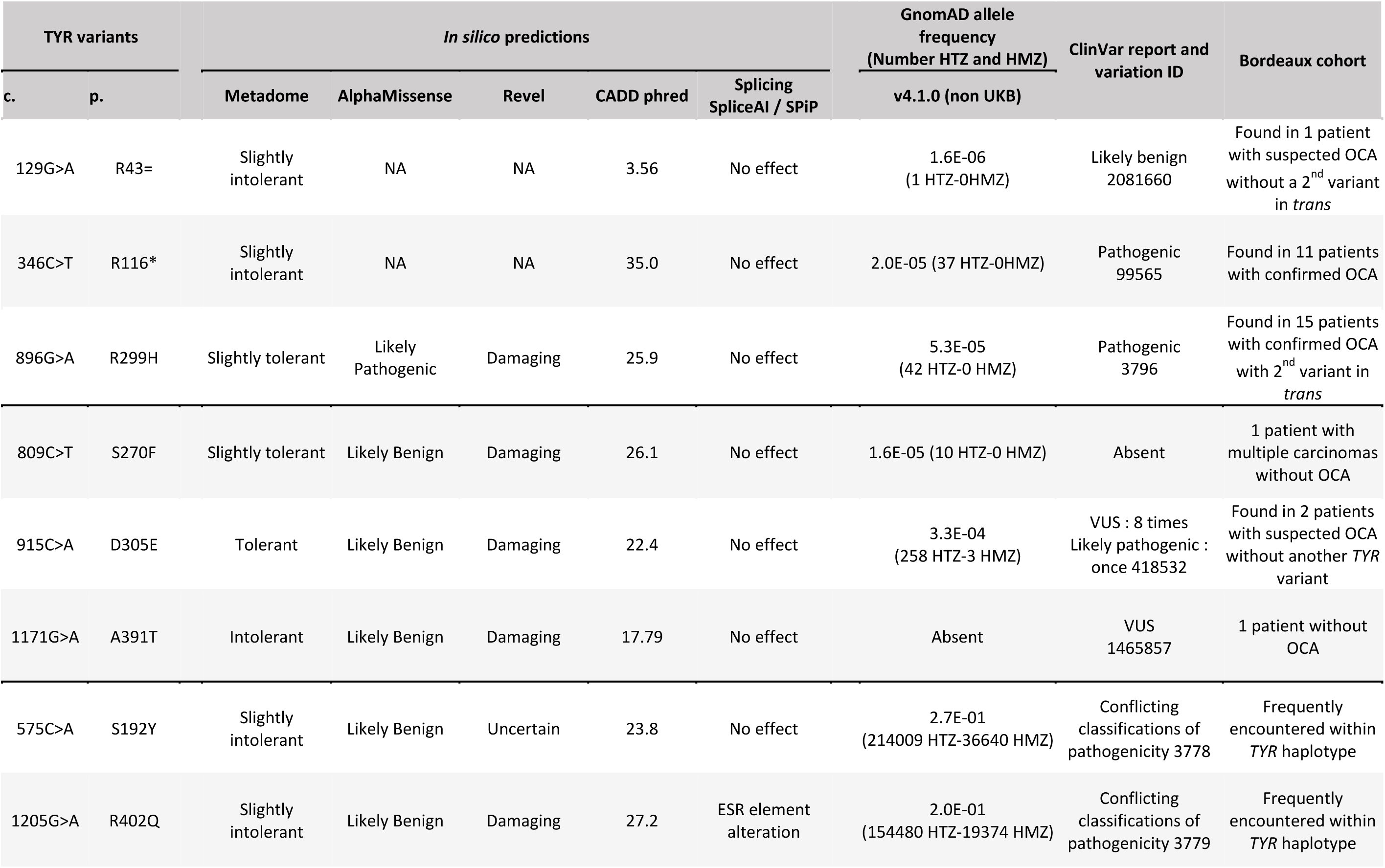
*In silico* predictions and allelic frequencies of variants included in this study. Nomenclature based on NM_000372.5. Predictions and frequency gathered from MobiDetails https://mobidetails.iurc.montp.inserm.fr/MD/, Splice AI : https://spliceailookup.broadinstitute.org/, SPiP : https://github.com/raphaelleman/SPiP, CADD : https://cadd.gs.washington.edu/snv, gnomAD : https://gnomad.broadinstitute.org/ and Clinvar : https://www.ncbi.nlm.nih.gov/clinvar/. Abbreviations: ESR, exonic splicing regulatory; HMZ, homozygous; HTZ, heterozygous; na, non-applicable; UKB, UK Biobank; VUS, variant of unknown significance.

| TYR variants |  | *In silico* predictions |  |  |  |  | GnomAD allele frequency<br>(Number HTZ and HMZ)<br>v4.1.0 (non UKB) | ClinVar report and<br>variation ID | Bordeaux cohort |
| --- | --- | --- | --- | --- | --- | --- | --- | --- | --- |
| c. | p. | Metadome | AlphaMissense | Revel | CADD phred | Splicing<br>SpliceAI / SPiP |  |  |  |
| 129G>A | R43= | Slightly intolerant | NA | NA | 3.56 | No effect | 1.6E-06<br>(1 HTZ-0HMZ) | Likely benign<br>2081660 | Found in 1 patient with suspected OCA without a 2<sup>nd</sup> variant in *trans* |
| 346C>T | R116* | Slightly intolerant | NA | NA | 35.0 | No effect | 2.0E-05 (37 HTZ-0HMZ) | Pathogenic<br>99565 | Found in 11 patients with confirmed OCA |
| 896G>A | R299H | Slightly tolerant | Likely Pathogenic | Damaging | 25.9 | No effect | 5.3E-05<br>(42 HTZ-0 HMZ) | Pathogenic<br>3796 | Found in 15 patients with confirmed OCA with 2<sup>nd</sup> variant in *trans* |
| 809C>T | S270F | Slightly tolerant | Likely Benign | Damaging | 26.1 | No effect | 1.6E-05 (10 HTZ-0 HMZ) | Absent | 1 patient with multiple carcinomas without OCA |
| 915C>A | D305E | Tolerant | Likely Benign | Damaging | 22.4 | No effect | 3.3E-04<br>(258 HTZ-3 HMZ) | VUS : 8 times<br>Likely pathogenic :<br>once 418532 | Found in 2 patients with suspected OCA without another TYR variant |
| 1171G>A | A391T | Intolerant | Likely Benign | Damaging | 17.79 | No effect | Absent | VUS<br>1465857 | 1 patient without OCA |
| 575C>A | S192Y | Slightly intolerant | Likely Benign | Uncertain | 23.8 | No effect | 2.7E-01<br>(214009 HTZ-36640 HMZ) | Conflicting<br>classifications of<br>pathogenicity 3778 | Frequently encountered within TYR haplotype |
| 1205G>A | R402Q | Slightly intolerant | Likely Benign | Damaging | 27.2 | ESR element alteration | 2.0E-01<br>(154480 HTZ-19374 HMZ) | Conflicting<br>classifications of<br>pathogenicity 3779 | Frequently encountered within TYR haplotype |

### 2.3 Rescue assay of missense VUS using a*TYR*-KO MNT1 cells

Four days after transfection with the recombinant expression vector containing the WT *TYR* cDNA, the rescue of melanin synthesis in *TYR*-KO MNT1 cells was clearly visible under the microscope. As shown in Fig. 2a, the transfected cells displayed densely pigmented melanosomes that were evenly distributed throughout their cytoplasm, including their dendrites. Around 20% of the cells were pigmented reflecting the usual efficacy of transient transfection in MNT1, and the level of pigmentation was consistent among the melanogenic cells. The assay was run for all variants and controls in parallel. There were visible differences in pigmentation depending on the variant carried by the recombinant expression vector. Dosage of *TYR* mRNA was performed by RT-qPCR showing a comparable level of transcription resulting from the expression of the WT and missense variant-carrying cDNAs (Fig. 2b). The tyrosinase was quantified by Western-blot (Fig. 2c). Although tyrosinase was detected in all samples derived from transfection with a cDNA carrying a missense variant, the amounts of protein significantly varied depending on the substitution. Variant D305E did not significantly affect the rate of tyrosinase expression. Variants A391T and S192Y resulted in lower tyrosinase levels (40-60% compared to WT). Variant R402Q alone led to reduced levels of tyrosinase expression at around 20% compared to controls. When combined to S192Y, variant R402Q resulted in a further reduction in tyrosinase content equivalent to the pathogenic control R299H with only 10% of residual signal. VUS S270F resulted in the same near to complete loss of tyrosinase detection. The corresponding cell pellet was homogeneously white as were the ones derived from transfection with pathogenic variants R116*, R299H, or the empty vector indicating that the S270F substitution impaired the rescue of pigment production (Fig. 2d). By contrast, cell pellets obtained after transfection with a *TYR* cDNA vector carrying VUS D305E were heavily pigmented as were the control pellets (WT and R43=). Comparative pigment dosage was performed on all samples by evaluating the absorbance of the solubilized tested extracts relative to the absorbance of the sample prepared from empty vector transfected cells. Absorbance was first monitored along a spectrum from 300 to 600 nm (Fig. S1) based on previous spectroscopic measurements performed on human skin melanin (18). Relative quantification was performed after reading the absorbance values at a chosen optimal wavelength of 400 nm (Fig. 2e). This confirmed the absence of melanin in samples derived from transfection with *TYR* cDNA carrying VUS S270F. Neither the two other rare VUS D305E and A391T nor S192Y, significantly impaired pigment production when compared to WT control or benign variant R43=. Significant reduction of melanin content was detected in rescue assays involving R402Q alone (∼37% compared to control). When combined in *cis* with S192Y the reduction of melanin content was even more pronounced with less than 15% of melanin concentration compared to controls. Microscopic observation of the transfected cells (Fig. S2) confirmed the complete absence of detectable pigment in rescue assays performed with variant S270F whereas cells with pigmented melanosomes were identified with D305E, A391T, S192Y and R402Q. Pigment was also visualized in a few cells transfected with the cDNA carrying S192Y and R402Q in *cis*. The distribution of this pigment was however mainly diffused with very few identified pigmented melanosomes compared to the positive controls. These results demonstrate an additive effect of the two frequent variants in a functional assay and suggest that the *cis* combination of S192Y and R402Q triggers endoplasmic reticulum retention of the mutated tyrosinase.

**Fig. 2:**
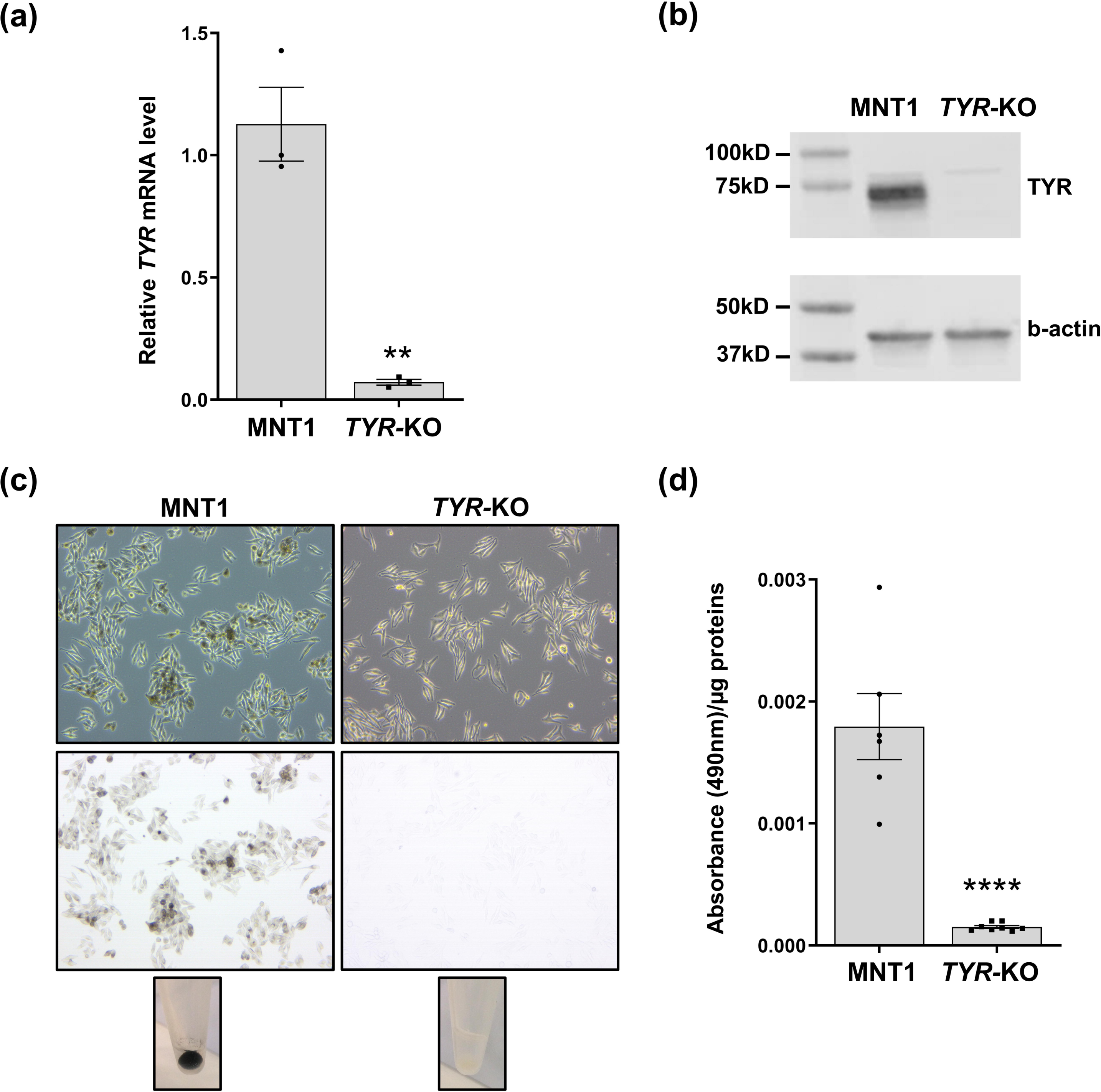
Rescue assay using selected variants and the *TYR*-KO MNT1 cell line. (a) Bright-field microscopy showing rescued pigmentation in two out of three *TYR-*KO cells transfected with control *TYR* cDNA (WT). Scale bar: 10µm. Functional rescue results in pigmented melanosomes evenly distributed in the cytoplasm including the dendrites (higher-magnification inset). (b) RT-qPCR analysis of *TYR* expression in *TYR-KO* transfected cells. *TYR* expression for each condition was normalized to the expression of *B2M* and compared to the WT condition using the 2^−ΔΔCt^ method. Data are presented as mean ± SEM (n=4). Kruskal Wallis test was performed and * indicates p<0.03. (c) Dosage of TYR protein expression by Western-blot. Quantification performed on three independent experiments is shown next to a representative Western-blot image. Tyrosinase signal intensities were normalized to b-actin. (d) *TYR-*KO cell pellets transfected with pCS2+ vector containing *TYR* cDNA with selected variants. (e) Melanin absorbance at 400nm for each condition is deduced from the background obtained with cells transfected with the empty vector before being normalized with total protein. Data are shown as mean ± SEM (n=4). For TYR protein and melanin dosage, test-values were compared to WT control by a one-way ANOVA followed by the Bonferroni post hoc multiple comparison test, **** indicates p<0.0001, *** indicates p<0.001, ** indicates p<0.01. Results in this Figure were obtained with the clone *TYR-*KO bearing the mutation c.879del/p.(E294Rfs*25).

## 3. Discussion

Several tyrosinase cell-based rescue assays have been used in a total of three publications to date (19–21). They are mostly using non-pigmentary cell types, such as the HEK293T embryonic renal cell line. Since these cells are devoid of melanosomes and lack the essential melanogenic enzymes TYRP1 and DCT, absorbance measurements of cell samples after rescue quantify the spontaneously oxidated products of dopachrome rather than melanin. While this measurement can be equated to a tyrosinase enzyme activity assay, it does not enable the targeting of the protein to the melanosome to be assessed. To approximate a pigmentary cell model, Li et al. suggested using the human melanoma-derived A375 cell line for the rescue test (19). A375 cells are amelanogenic in culture. However, the causes of this cell line’s inability to pigment are complex. They appear to be linked to low catalase expression, which leads to inhibition of endogenous TYR, TYRP1 and DCT enzymes (22). The rescue test proposed by Li et al. in A375 cells also requires the addition of excess tyrosine, which may introduce bias when interpreting the pigment levels quantitatively. MNT1 cells are also derived from human melanoma but resemble primary melanocytes in many aspects (transcriptome, morphology, melanosomal content). Contrary to A375 cells, they do express the three melanogenic enzymes that locate at the melanosomal membrane. MNT1 cells produce final melanin that is eventually secreted in the form of melanocores (10).

Some previously identified variants of *TYR* have been shown to impair glycosylation and cause endoplasmic retention of the tyrosinase. The most commonly described example is variant R402Q that has first been identified as associated with a mild form of albinism when in *trans* to a pathogenic variant (23). This has been confirmed by independent family studies (24). Functional studies indicated that this variant results in a temperature-dependent reduction of the enzymatic activity *in vitro* as well as when expressed in cultured HeLa cells (7,25,26). Interestingly, Jagirdar et al. reported lower tyrosinase content, trafficking and activity in homozygous R402Q primary melanocytes compared to wild-type cells (25). More recently, we found that variant R402Q is significantly associated with albinism when in *cis* to c.-301C, which is located in the promoter of *TYR* and significantly impacts its activity (4). Interestingly, the homozygote haplotype [c.-301C; R402Q] is associated with albinism. Variant S192Y seems to have an additional effect on [c-301C; R402Q]. However, the study was unable to assess such an effect of S192Y on a [c-301C>T; R402Q] haplotype due to significant linkage disequilibrium between S192Y and c.-301C, which made the corresponding diplotypes unavailable for statistical analysis. Our functional assay confirms the presence of an additive effect of S192Y in *cis* to R402Q within a melanogenic system, which suggests that the haplotype [c.-301C>T; S192Y; R402Q] may be pathogenic. Future molecular diagnosis will consider the three alleles in order to verify this hypothesis.

Three VUS were tested in this study. We show that the rare variant S270F (allele frequency =1.6 10^-5^) is pathogenic as it fails to rescue pigment production in the assay. The two other rare missense variants, D305E and A391T did not prevent rescue of pigmentation. However, A391T displayed a profile similar to S192Y with significantly reduced tyrosinase expression and slightly lower pigment rescue than the controls. This suggests that A391T is likely benign on its own. However, an additive effect in *cis* with hypomorphic variants such as S192Y, R402Q or others will be worth considering in future studies, with the perspective of possibly identifying a pathogenic haplotype to be considered in the context of diagnosis.

Targeting pigmentation genes in the heavily pigmented MNT1 cell line is straightforward as clones of interest can be selected by direct observation under a bright field microscope. As we show here, these clones can be used in a simple rescue assay for testing variants of unknown significance. The same strategy will be worth applying to variants in other albinism genes given the fact that they require cells to express functional tyrosinase. We have already engineered KO clones targeting *TYRP1* (OCA3) and *SLC45A2* (OCA4) and find that they each display a directly observable discontinuous phenotype (Fig. S3). *SLC45A4*-KO MNT1 clones are completely devoid of pigment reflecting the epistatic interaction between *SLC45A2* and *TYR*. *TYRP1*-KO MNT1 clones are devoid of black melanin making their pigmentation uniformly brown. These clones can therefore serve as a host system for respectively testing missense VUS of *SLC45A2* (204 reports in ClinVar) and *TYRP1* (246 reports in ClinVar).

To conclude, we provide a new straightforward cell-based assay for functionally evaluating yet unclassified variants in albinism genes which should prove useful in the context of molecular diagnosis.

## 4. Materials and Methods

### 4.1 Gene editing

The *TYR* gene was knocked out by the CRISPR/Cas9 gene editing system in MNT1 cells (ATCC CRL-3450). The guide RNA (gRNA) (5’ TTATGCAATGGAACGCCCGA 3’) that targets exon 2 was selected using the Custom Alt-R CRISPR-Cas9 gRNA web resources (Integrated DNA Technologies). The selected gRNA was tested with the Crispor program, UC Santa Cruz (27) for a specificity prediction score >95, and minimal number of off-target loci. The day before transfection, 1×10^5^ MNT1 cells were seeded in a 12-well plate and cultured in DMEM+GlutaMAX (Gibco) supplemented with 1% penicillin and streptomycin, 1% non-essential amino acids and 10% fetal bovine serum (Gibco). Transfection was performed using the TrueCut Cas9 Protein v2 kit with 20pmol of Cas9 Protein v2, 20pmol of gRNA, 6µL of Lipofectamine Cas9 Plus Reagent and 3.6µL of Lipofectamine CRISPRMAX according to the manufacturer’s instructions (Invitrogen). Forty-eight hours after transfection, genomic DNA from a 1×10^4^ cells aliquot was analyzed by PCR and sequencing. DNA was extracted and amplified using Phire Tissue Direct PCR Master Mix (Thermofisher) according to the manufacturer’s instructions. The targeted sequence was amplified by PCR (forward primer: 5’-ACTCAGCCCAGCATCATTCT-3’; reverse primer: 5’-TTGAAGAGGACGGTGCCTT-3’) for 30 cycles (98°C for 5s, 58°C for 5 s, 72°C for 20s). PCR products were analyzed by Sanger sequencing (Eurofins). The TIDE online resource was used to verify that targeting efficiency was higher than 15% (28). *TYR-*KO cell clones were isolated by limiting dilution of 0.8 cell per well. After 4 days, wells with no colonies or more than one colony were removed from the screening. Single colonies were cultured until non-pigmented *TYR-*KO clones could be discriminated from pigment producing cells resulting in selection of ∼10% of clones. Selected clones were analyzed by sequencing as above.

### 4.2 Selected *TYR*-KO clones

Three independent homozygote clones each carrying a frameshift variant leading to a premature stop codon were selected for this study. The corresponding mutations are: c.877dup/p.(E294Rfs19*), c.878_879del/p.(P293Rfs17*) and c.879del/p.(E294Rfs*25). The clone bearing the c.879del/p.(E294Rfs*25) variant was used to design the rescue assay.

### 4.3 Recombinant plasmid construct and site-directed mutagenesis

Total RNAs from SK-MEL-3 cells (ATCC HTB-69) were purified with the RNeasy Mini kit (Qiagen) according to manufacturer’s recommendations. Reverse transcription (RT) was performed by oligo(T)-primed reverse-transcription using M-MLV reverse transcriptase (Promega) with 1µg of RNAs. *TYR* cDNA was amplified with the Q5 High Fidelity PCR kit (New England Biolabs) according to the manufacturer’s instructions with the following primers (forward: 5’-AATGCTCCTGGCTGTTTTGT-3’; reverse: 5’-TGGCCCTACTCTATTGCCTAA-3’) and purified using the Monarch PCR & DNA Cleanup kit (New England Biolabs). Cloning in the pGEM-T Easy Vector Systems (Promega) was performed in a 1:1 ratio (insert:vector) following the manufacturer’s instructions. From a positive clone, *TYR* cDNA was PCR-amplified with CloneAmp HiFi PCR Premix (TakaroBio) according to manufacturer’s instructions using the following primers (forward : 5’-CCATCGATTCGAATTAATGCTCCTGGCTGTTTTGT-3’; reverse: 5’-GAGAGGCCTTGAATTTGGCCCTACTCTATTGCCTAA-3’) and treated with the In-Fusion HD Cloning kit with Cloning Enhancer Treatment protocol (TakaraBio) before being recombined with EcoRI linearized pCS2+ vector (GeneCust). The resulting cDNA was sequenced and the presence of two frequent variants (c.575C>A/p.S192Y and c.824T>C/p.V275A) was noted. Using the QuickChange Site-Directed Mutagenesis kit (Agilent) following the manufacturer’s recommendations, variant c.824T>C/p.V275A was removed from the cloned cDNA which resulted in the S192Y carrying vector. Subsequently, from S192Y vector, c.575C>A/p.S192Y variant was removed to produce the WT cDNA. All constructs carrying single VUS, a nonsense and a missense pathogenic variant (as rescue negative controls) and one benign variant (as rescue positive control) were derived from the WT vector (variants listed in table S1). The vector containing S192Y and R402Q was obtained by introduction the R402Q mutation in the S192Y bearing construct. All primers for targeted mutagenesis were designed as recommended (Agilent) and are listed in Table S1. All mutated vectors were confirmed by Sanger sequencing (Eurofins).

### 4.4 Cell transient transfection

*TYR-*KO cells were cultured in DMEM+GlutaMAX (Gibco) supplemented with 1% penicillin and streptomycin, 1% non-essential amino acids and 10% fetal bovine serum (Gibco). Two days before transfection cells were seeded in 6-well plates with 3×10^5^ cells per well. Transfections were performed using Lipofectamine3000 (Thermo Fisher Scientific) with 2.5µg vector according to optimized manufacturer’s instructions. Optimization of the standard protocol consisted in increasing the mix incubation time from 5 to 20 minutes. To improve cell transfection efficiency, the transfection mix was applied to the cells in fresh Opti-MEM 1X (Gibco). The cells were left in Opti-MEM at 37°C for 4 hours after which it was replaced by complete culture medium. Cells were collected 96 hours after transfection by scraping in PBS. For each well, cells were divided into 2 dry pellet samples (one for melanin dosage and Western-blot and one for RT-qPCR) that were stored at -80°C.

### 4.5 RT-qPCR expression measurement

Total RNAs were purified using the RNeasy Micro kit including DNase treatment (Qiagen) from 1×10^5^ cells. First-strand cDNA synthesis was carried out with ∼1µg RNA as described before. The RT products are then diluted by ½. All real-time RT-PCRs were carried out using the iQ™ SYBR Green Supermix 2X (BioRad) in a final volume of 25 µl with 200nM of primers and 2µL of diluted cDNA following the manufacturer’s protocol. Primers selected target exons 2 and 3 of *TYR* (forward: 5’-TACGGCGTAATCCTGGAAAC-3’; reverse: 5’-ATTGTGCATGCTGCTTTGAG-3’) as well as *β2 microglobulin (B2M)* (forward: 5’-GGATCGAGACATGTAAGCA-3’; reverse: 5’-CAATCCAAATGCGGCATCT-3’). The qPCR reaction performed using a QuantStudio3 thermocycler (Applied Biosystems) included an initial denaturation step at 95°C for 3 minutes followed by 40 cycles of 95°C for 15s and combined annealing and extension at 60°C for 30s with standard temperature ramping mode (1.6°C/s). Fluorescence was monitored at the end of each cycle and data were analyzed using the QuantStudio design and analysis software (2.7.0).

### 4.6 Melanin measurement

The analysis of pigment loss in the three *TYR-*KO clones was initially performed using a conventional melanin assay method as described (29). Melanin dosage was then optimized for the quantitative pigment rescue assay. Sample pellets were resuspended in 50µL of Pierce RIPA buffer (ThermoFisher Scientific) supplemented with 1% of protease inhibitors cocktail (Sigma-Aldrich). The samples were vortexed, agitated for 10 minutes at 4°C and centrifuged at 4°C for 10 minutes at 16 000g. The supernatant was kept at 4°C for protein assay and Western-blot. The pellet used for melanin dosage was dissolved 10 minutes at 100°C in 50µL of 1N NaOH and left to cool to room temperature. Twenty µL of each sample were loaded in duplicate into a 384-well UV-STAR plate (Greiner). The absorbance was measured on a spectrum extending from 300nm to 600nm using a CLARIOstar Plus spectrophotometer (BMG Labtech). Total protein quantification was performed using ¼ dilution of the collected supernatants, according to the instructions of the Pierce BCA Protein Assay kit (ThermoFisher Scientific). Absorbance was measured at 570nm in a 96-well plate using a Safas MP96 spectrophotometer. For each sample, melanin dosage was estimated from the sample absorbance at 400nm normalized by the absorbance measured in samples from cells transfected with empty pCS2+. This concentration was then normalized relative to the amount of total protein.

### 4.7 Western-Blot

Protein extracts were denatured at 95°C for 5 minutes with 1X Laemmli (Biorad) and 10% of 2-mercaptoethanol. Denatured proteins (20µg) were loaded on a 4–20% Mini-PROTEAN TGX Precast Protein Gel and migrated in 1X Tris/Glycine/SDS buffer (Biorad). Proteins were transferred on Trans-Blot Turbo Midi Nitrocellulose membrane (Biorad) with 1X Trans-Blot transfer buffer (Biorad) using the Trans-Blot Turbo Transfer System (Biorad). The membrane was saturated with Intercept PBS blocking buffer (LI-COR) for 45 minutes under agitation at room temperature. Tyrosinase and b-actin proteins were revealed sequentially with a specific mouse monoclonal antibody. The membrane was first incubated with a solution containing blocking buffer, 0.1% Tween and anti-tyrosinase antibody (1/600, T311, Merck) and left overnight at 4°C with agitation before being incubated with IRDye 800CW Goat anti-mouse antibody (1/5000, LI-COR) in blocking buffer mixed with Tween 0,05%/PBS 1X (v/v) for 45 minutes under agitation. The tyrosinase signal was revealed with the OdysseyXF (LI-COR) after ∼2 minutes of exposure. The membrane was then incubated under the same conditions with an anti-b-actin antibody (1/5000, AC-15, Merck) for 45 minutes at room temperature and exposed in the Odyssey apparatus for ∼30 sec. Each band signal was quantified by the ImageJ software (https://imagej.net/ij/). Relative tyrosinase levels were deduced after normalization with b-actin.

### 4.8 Statistical analysis

All the results are expressed as mean value ± standard error of the mean (SEM) of at least 3 independent experiments, unless otherwise stated. Statistical analysis was performed with GraphPad Prism version 8.0.2 for Windows using Student’s t test, one-way ANOVA followed by the Bonferroni post hoc multiple comparison test or Kruskal Wallis test.

### 4.9 Data availability

Variants have been uploaded to ClinVar public database.

## Acknowledgments

We are very grateful to Genespoir (the French albinism association) for their support and the significant help they provide to patients. We thank Elodie Muzotte for her assistance with bright-field microscopy.

## Legends to Figures and Tables

**Fig. S1: Representative absorbance spectrum obtained during melanin assay of selected VUS.**

The absorbance was measured on a spectrum extending from 300nm to 600nm using a CLARIOstar Plus spectrophotometer (BMG Labtech)

**Fig. S2: Bright-field microscopy images of *TYR*-KO MNT1 cells after rescue assay.**

For each construct, a representative image is provided (bar scale :20µm). No pigmented cells were identified after transfection with recombinant plasmids containing R116*, R299H and S270F. In the rescue assay with S192Y;R402Q, pigment was mostly diffuse.

**Fig. S3: Loss of function of *TYRP1* and *SLC45A2* genes in MNT1 cells.**

(a) Loss of function of the *TYRP1* and *SLC45A2* genes in MNT1 cells results in a marked decrease or abolition of pigmentation as observed by contrast and bright field microscopy. Magnification x40. (b) Loss of function of the *TYRP1* gene leads to a change in pigment composition as observed by microscopy at high magnification (x400).

**Table S1: List of primers used for site-directed mutagenesis.**

Primer pairs used to reverse variants carried by the ancestral *TYR* cDNA cloned from the SK-MEL-3 cell line are indicated with ^$^.

## Abbreviations

ANOVA: ANalysis Of Variance
cDNA: complementary DNA
CRISPR: Clustered Regularly Interspaced Short Palindromic Repeats
gnomAD: Genome Aggregation Database
gRNA: guide RNA
KO: Knock Out
mRNA: messenger RNA
NMD: Nonsense-Mediated Decay
OCA1: Oculo-Cutaneous Albinism type 1
OCA3: Oculo-Cutaneous Albinism type 3
OCA4: Oculo-Cutaneous Albinism type 4
RT: Reverse Transcription
RT-qPCR: Reverse Transcriptase quantitative Polymerase Chain Reaction
SEM: Standard Error of Mean
SNV: Single Nucleotide Variant
VUS: Variants of Unknown Significance
WT: Wild Type

