## Supplementary figures and images for "A new rescue assay for genetic diagnosis of oculocutaneous albinism using MNT1 knock-out cells"

### Fig. S1

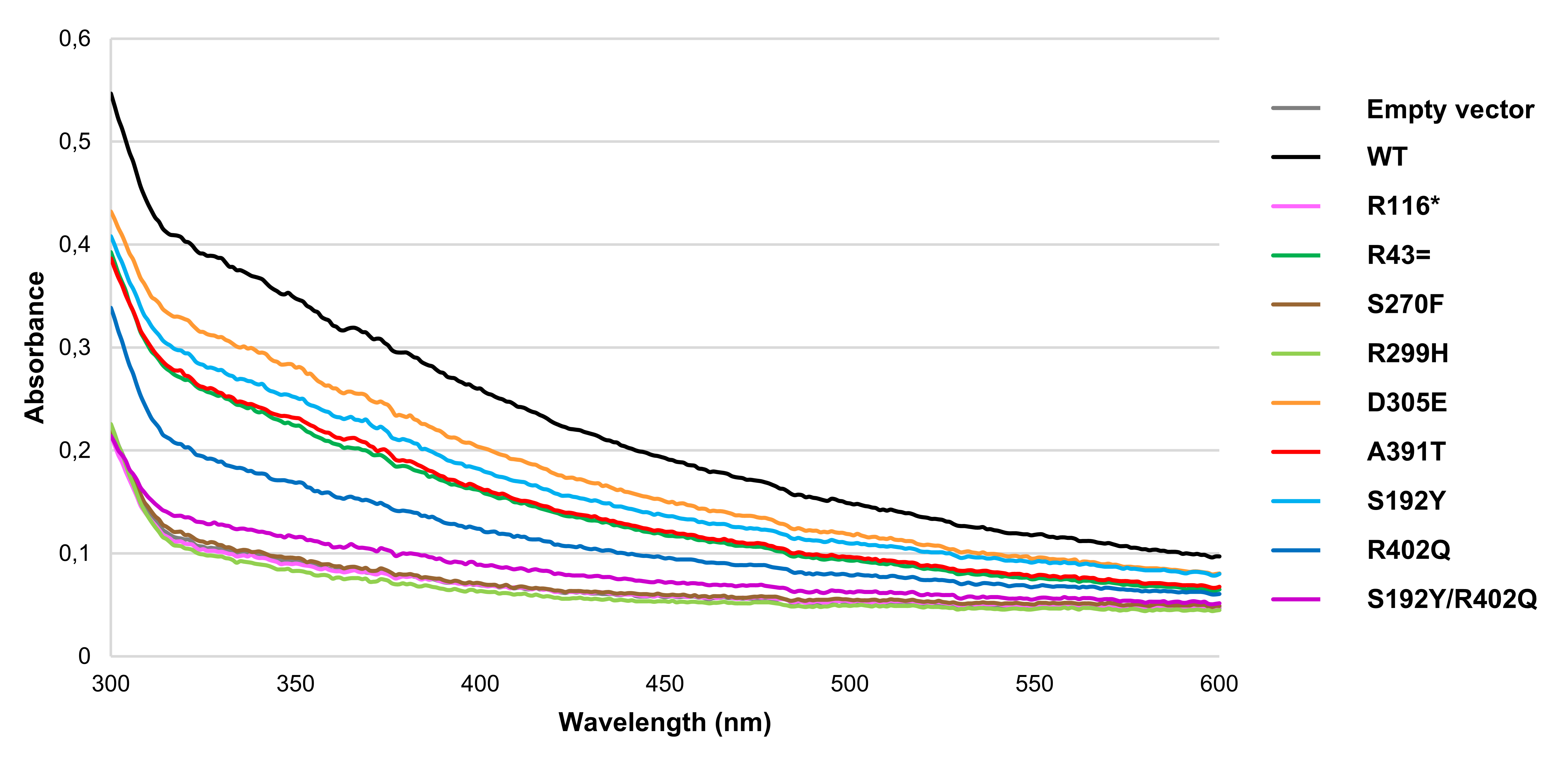

### Fig. S2

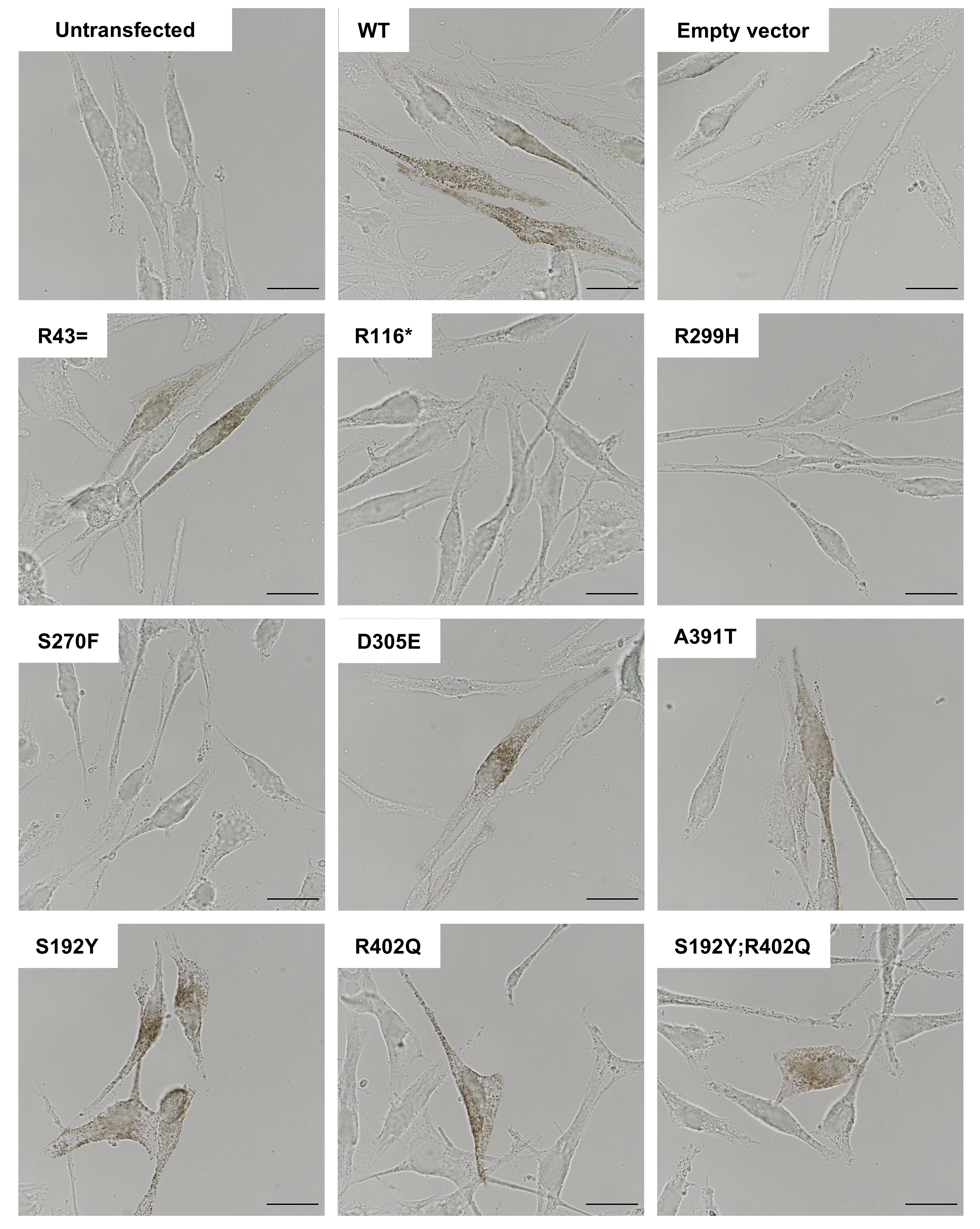

### Fig. S3

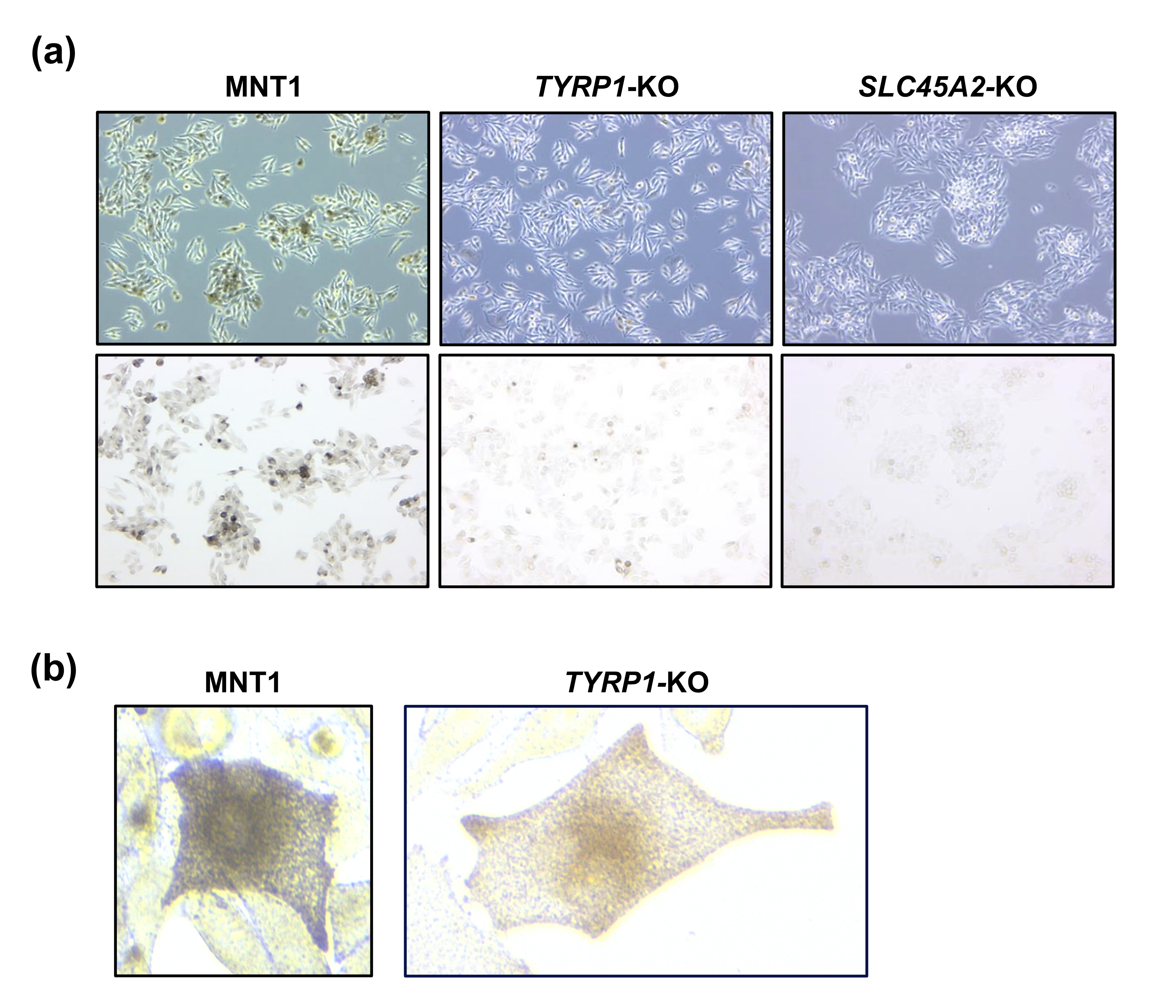
