## Supplementary material for "A new rescue assay for genetic diagnosis of oculocutaneous albinism using MNT1 knock-out cells": Table S1

| c.129G>A/p.R43= | Forward : 5’- GTGGAGCGGGGACAG**A**AGTCCCTGTGGCCAG -3’ |
| --- | --- |
|  | Reverse : 5’- CTGGCCACAGGGACT**T**CTGTCCCCGCTCCAC-3’ |
| c.346C>T/p.R116* | Forward : 5’-CGGGACCAAACTGCACAGAGAGA**T**GACTCTTGGTGAGAAG-3’ |
|  | Reverse : 5’-CTTCTCACCAAGAGTC**A**TCTCTCTGTGCAGTTTGGTCCCC-3’ |
| **^$^** c.575A>C/p.Y192S | Forward : 5’-GCACTGCTTGGGGGAT**C**TGAAATCTGGAGAGAC-3’ |
|  | Reverse : 5’-GTCTCTCCAGATTTCA**G**ATCCCCCAAGCAGTGC-3’ |
| c.809C>T/p.S270F | Forward : 5’-CTTACTCAGCCCAGCAT**T**ATTCTTCTCCTCTTGGC-3’ |
|  | Reverse : 5’-GCCAAGAGGAGAAGAAT**A**ATGCTGGGCTGAGTAAG-3’ |
| **^$^** c.824C>T/p.A275V | Forward : 5’-CCTCTTGGCAGATTGT**C**TGTAGCCGATTGGAG-3’ |
|  | Reverse : 5’-CTCCAATCGGCTACA**G**ACAATCTGCCAAGAGG-3’ |
| c.896G>A/p.R299H | Forward : 5’-CGAGGGACCTTTACGGC**A**TAATCCTGGAAACC -3’ |
|  | Reverse : 5’-GGTTTCCAGGATTA**T**GCCGTAAAGGTCCCTCG-3’ |
| c.915C>A/p.D305E | Forward : 5’-AATCCTGGAAACCATGA**A**AAATCCAGAACCCCAGG-3’ |
|  | Reverse : 5’-CCTGGGGTTCTGGATTT**T**TCATGGTTTCCAGGATT -3’ |
| c.1171G>A/p.A391T | Forward :  5’-CCTTCTTCACCAT**A**CATTTGTTGACAGTATTTTTGAGCAGTGGC-3’ |
|  | Reverse :  5’-GCCACTGCTCAAAAATACTGTCAACAAATG**T**ATGGTGAAGAAGG-3’ |
| c.1205G>A/p.R402Q | Forward : 5’-TTGAGCAGTGGCTCC**A**AAGGCACCGTCCTCT-3’ |
|  | Reverse : 5’-AGAGGACGGTGCCTT**T**GGAGCCACTGCTCAA-3’ |
